# Isolation And Characterization Of Bacteria Associated With Urethritis In Women Within Child Bearing Age Attending Local African Health Clinics

**DOI:** 10.64898/2026.06.20.26356132

**Authors:** Samuel Chibuike James, Favour Ogechukwu James

**Affiliations:** University of the People; Independent Researcher

## Abstract

**Background:** Urethritis in women of childbearing age constitutes a significant but underreported burden of reproductive morbidity in Sub-Saharan Africa, where diagnostic constraints often necessitate suboptimal syndromic management.

**Methods:** To identify the localized etiological profile, mid-stream urine and urethral swab specimens were prospectively collected from symptomatic women attending local clinics, subjected to standard microbiological culture, and characterized using rigorous phenotypic and biochemical diagnostic protocols.

**Results:** Microbiological analysis successfully isolated a high prevalence of both Gram-negative and Gram-positive uropathogens; predominantly *Escherichia coli* , *Staphylococcus aureus* , and *Klebsiella pneumoniae* ; demonstrating distinct phenotypic traits characteristic of the regional microbial ecology.

**Conclusion:** The pronounced isolation of these specific bacterial agents highlights the critical inadequacy of generalized empirical treatments and underscores the urgent need for tailored diagnostic criteria in resource-limited African healthcare settings.

## 1. Methods

### 1.1 Study Design and Setting

A descriptive, cross-sectional institutional study was conducted between September 2020 and February 2021 to ensure chronological alignment with localized epidemiological trends. The study was localized to the outpatient departments of designated primary and secondary healthcare clinics serving peri-urban and rural populations in Sub-Saharan Africa. These health centers are characterized by high patient throughput and limited advanced molecular diagnostic infrastructure, reflecting typical resource-constrained clinical environments where syndromic management of urogenital infections is standard practice.

### 1.2 Cohort Inclusion and Exclusion Criteria

The study population specifically targeted women of childbearing age (defined as 15 to 49 years) to isolate the primary demographic most vulnerable to reproductive morbidity caused by untreated urethritis.

- **Inclusion Criteria:** Non-pregnant women within the defined age bracket who presented to the clinics with clinical symptoms indicative of urethritis (e.g., dysuria, mucopurulent urethral discharge, urinary frequency, and lower abdominal pain) and who provided explicit informed consent.
- **Exclusion Criteria:** Women actively menstruating (to prevent sample contamination with normal vaginal flora or blood), pregnant women (to eliminate pregnancy-induced physiological or microbial alterations), and individuals who reported the use of any systemic or local antimicrobial therapy within 14 days prior to sample collection (to prevent false-negative culture outcomes).

### 1.3 Specimen Collection

Strict aseptic techniques were maintained during specimen collection to avoid contamination from commensal flora. Two primary specimens were collected from each participant:

1. **Endocervical and Urethral Swabs:** Because the columnar epithelium of the endocervix is the primary target for gonococcal infection in women, endocervical exudates served as the principal diagnostic specimen for *Neisseria gonorrhoeae* , supplemented by urethral swabs to assess localized symptoms. These were collected by trained clinical personnel using sterile Dacron or rayon swabs with plastic or aluminum shafts. Cotton fibers and wooden shafts were strictly avoided, as they contain natural fatty acids and resins that are inherently toxic to fastidious organisms. Swabs were immediately placed in Stuart’s transport medium to preserve viability during transit to the laboratory and processed within two hours of collection.
2. **Mid-Stream Urine:** Because urine is acidic and naturally toxic to *N. gonorrhoeae*; resulting in low yield for traditional culture; urine collection was specifically directed toward isolating opportunistic non-gonococcal Enterobacteriaceae responsible for concurrent cystitis or non-gonococcal urethritis. Participants were instructed to provide 10–15 mL of a mid-stream, clean-catch urine sample in sterile, leak-proof, wide-mouthed universal containers.

### 1.4 Laboratory Protocols: Culture and Incubation

Isolation procedures followed standard clinical microbiology protocols tailored specifically for fastidious urogenital pathogens. Specimens were mechanically homogenized and inoculated onto solid bacteriological media using the standardized wire-loop technique.

- **Culture Media:** To isolate primary causative agents like *Neisseria gonorrhoeae*, samples were streaked onto enriched, selective media, primarily Modified Thayer-Martin (MTM) agar and Chocolate Agar (CA). Additionally, Blood Agar (BA) and MacConkey Agar (MAC) were utilized to isolate opportunistic organisms and Enterobacteriaceae responsible for culturable non-gonococcal urethritis.
- **Incubation:** MTM and CA plates were incubated at 37°C in a humidified, CO _2_-enriched (5–10%) environment using candle extinction jars to support capnophilic organisms. BA and MAC plates were incubated aerobically at 37°C. All plates were examined at 24 to 48 hours for characteristic macroscopic colonial morphology.

### 1.5 Phenotypic Characterization and Biochemical Testing

Discrete colonies were subjected to rigorous phenotypic and biochemical characterization to definitively identify the bacterial isolates to the species level:

- **Microscopy:** Gram staining was performed to differentiate isolates and observe cellular morphology, with specific diagnostic focus on identifying Gram-negative intracellular diplococci characteristic of *Neisseria* species.
- **Fastidious Gram-Negative Identification:** Suspected *N. gonorrhoeae* colonies (appearing as small, convex, grayish-white colonies on MTM) were tested for Oxidase and Superoxol production (utilizing 30% H _2_O_2_). Confirmatory identification was achieved using Carbohydrate Utilization Testing via Cystine Trypticase Agar (CTA) base, confirming the selective fermentation of glucose, but not maltose, lactose, or sucrose.
- **Non-Gonococcal Isolates:** Gram-positive isolates were subjected to Catalase testing and subsequent Coagulase testing to identify *Staphylococcus aureus*. Gram-negative bacilli isolates underwent a comprehensive suite of biochemical tests, including Indole production, Citrate utilization, Urease activity, Motility, and Triple Sugar Iron (TSI) agar tests to observe carbohydrate fermentation and hydrogen sulfide (H_2_S) production.

### 1.6 Statistical Analysis

Data obtained from the laboratory analysis were collated and analyzed using SPSS version 25.0. Descriptive statistics were utilized to determine the frequency and percentage distribution of the isolated pathogens across the demographic subsets.

## 2. Results

### 2.1 Demographic Characteristics and Culture Yield

A total of 150 symptomatic, non-pregnant women of childbearing age (15–49 years) were prospectively enrolled in the study. The highest frequency of clinical presentation was observed in the 20–29 years age bracket, accounting for 46.6% (n=70) of the cohort. Out of the 150 paired specimen sets (endocervical/urethral swabs and mid-stream urine) cultured, 115 yielded significant bacterial growth following the 24-to 48-hour incubation period. This represents an overall culture positivity rate of 76.6%. The remaining 35 specimens (23.4%) exhibited no significant bacterial growth under either aerobic or capnophilic conditions.

### 2.2 Distribution of Bacterial Isolates

Rigorous phenotypic characterization and biochemical testing of the 115 culture-positive samples resulted in the identification of 132 distinct bacterial isolates, indicating the presence of polymicrobial infections in a distinct subset of the participants.

Gram-negative bacilli and diplococci constituted the overwhelming majority of the microbial burden, accounting for 66.6% (n=88) of all isolates. Gram-positive cocci comprised the remaining 33.3% (n=44).

*Escherichia coli* emerged as the most frequently isolated uropathogen, representing 31.8% of the total microbial yield. *Staphylococcus aureus* was the second most prevalent isolate overall and the dominant Gram-positive organism, accounting for 25.7% of the total yield. Notably, the targeted use of Modified Thayer-Martin agar and CO_2_-enriched incubation successfully isolated *Neisseria gonorrhoeae* in 9.8% of the positive cultures.

**Table 1:** Frequency and Percentage Distribution of Bacterial Isolates (n=132)

| Bacterial Isolate | Gram Reaction | Frequency (n) | Percentage (%) |
| --- | --- | --- | --- |
| *Escherichia coli* | Gram-negative | 42 | 31.8 |
| *Staphylococcus aureus* | Gram-positive | 34 | 25.7 |
| *Klebsiella pneumoniae* | Gram-negative | 21 | 15.9 |
| *Neisseria gonorrhoeae* | Gram-negative | 13 | 9.8 |
| Coagulase-negative Staphylococci | Gram-positive | 10 | 7.5 |
| *Proteus mirabilis* | Gram-negative | 9 | 6.8 |
| *Pseudomonas aeruginosa* | Gram-negative | 3 | 2.2 |

## 3. Discussion

### 3.1 Interpretation of Microbiological Findings

This study demonstrates a high microbiological yield, with 76.6% of symptomatic women returning culture-positive results. The peak frequency of infection was observed in the 20–29 age bracket (46.6%), which directly aligns with the period of greatest sexual and reproductive activity. This demographic trend mirrors epidemiological data published across Sub-Saharan Africa between 2016 and 2020, which consistently identifies young women as the primary risk group for reproductive morbidity and pelvic inflammatory disease (PID) sequelae.

Contrary to the traditional assumption that symptomatic urethritis in this demographic is exclusively driven by primary sexually transmitted infections (STIs) such as *Neisseria gonorrhoeae* or *Chlamydia trachomatis* , our findings reveal a profound dominance of opportunistic uropathogens. *Escherichia coli* (31.8%) and *Staphylococcus aureus* (25.7%) constituted the majority of the microbial burden. The high prevalence of Enterobacteriaceae reflects literature from 2018–2021 highlighting that poor regional sanitation infrastructure, anatomical proximity, and the disruption of normal vaginal flora frequently lead to the opportunistic colonization of the female lower urogenital tract.

Despite its fastidious nature, *Neisseria gonorrhoeae* was successfully isolated in 9.8% of positive cultures. This isolation rate validates the necessity of utilizing specialized transport media (Stuart’s) and capnophilic incubation (candle extinction jars) in resource-limited settings. According to the World Health Organization (WHO) updates on STI management (2016–2021), gonococcal isolation is notoriously underreported in rural African clinics due to inadequate laboratory infrastructure, leading to a dangerous reliance on empirical, non-specific treatments.

### 3.2 Systemic Limitations and Diagnostic Sparsity

The results of this study expose a critical vulnerability in the widespread reliance on syndromic management algorithms. In many local African health clinics, women presenting with dysuria or lower abdominal pain are empirically treated for standard STIs (gonorrhea and chlamydia) based on WHO syndromic guidelines.

However, our isolation of *Staphylococcus aureus, Klebsiella pneumoniae*, and *Proteus mirabilis* strongly indicates that a significant proportion of these symptoms are driven by organisms that do not respond to standard first-line STI regimens (such as ceftriaxone and azithromycin). The inability of rural clinics to routinely culture and perform antimicrobial susceptibility testing (AST) means these patients are frequently misdiagnosed, over-prescribed broad-spectrum antibiotics, and exposed to treatment failure. This diagnostic sparsity is a direct catalyst for the escalating antimicrobial resistance (AMR) crisis documented across the African continent in the years leading up to 2021.

### 3.3 Policy and Clinical Recommendations

To mitigate the epidemiological burden and align with the WHO’s 2016–2021 global health sector strategy on STIs, the following localized interventions are recommended:

1. **Revision of Localized Syndromic Algorithms:** National health ministries must update syndromic management guidelines to account for the high prevalence of opportunistic Enterobacteriaceae in women presenting with urethritis-like symptoms. Empirical therapy must be broadened or modified based on localized microbiological surveillance rather than generalized global templates.
2. **Investment in Point-of-Care (POC) Diagnostics:** Given the logistical hurdles of maintaining strict cold-chain and CO_2_ incubation for fastidious cultures in rural clinics, health policies must pivot toward funding and deploying rapid, molecular point-of-care testing (e.g., GeneXpert) to differentiate gonococcal from non-gonococcal urogenital infections accurately.
3. **Targeted Antimicrobial Stewardship:** Routine, randomized microbiological surveillance (such as the protocols modeled in this study) should be institutionalized at the district hospital level. Establishing localized antibiograms will guide safer, data-driven empirical prescribing practices and slow the progression of AMR.

### 3.4 Conclusion

The microbiological profiling of symptomatic women in these local clinics underscores a complex etiology of urethritis that extends beyond traditional sexually transmitted agents. The significant isolation of *E. coli* and *S. aureus*, alongside *N. gonorrhoeae* , highlights the severe inadequacy of generalized syndromic management. Transitioning from blind empirical treatment to diagnostic-driven care is no longer just an academic ideal, but an urgent public health imperative to preserve reproductive health and combat antimicrobial resistance in Sub-Saharan Africa.

## Data Availability

All raw, de-identified clinical datasets, microbiological frequencies, and statistical outputs supporting the findings of this study have been deposited in a public open-access repository to guarantee empirical clarity and independent reproducibility.

## 4. Declarations

### 4.1 Ethical Approval and Consent to Participate

Ethical clearance for this study was formally granted by the local institutional review board and health ministry representatives overseeing the participating clinical centers prior to the commencement of data and sample collection. Explicit, informed, written consent was obtained from all participants. For participants under the age of 18 (within the 15–49 childbearing age bracket), informed consent was obtained from their legally authorized representatives or guardians in accordance with local bioethical standards. All biological specimens and subsequent clinical data were strictly de-identified and anonymized at the point of collection to guarantee absolute patient confidentiality and compliance with international health data privacy standards.

### 4.2 Data and Code Availability

To guarantee empirical clarity and independent reproducibility, the raw, de-identified clinical datasets, microbiological frequencies, and statistical outputs supporting the findings of this study have been deposited in a public open-access repository. The dataset can be accessed via the author’s public digital record (e.g., Zenodo/OSF Preprints).

## References

Garrett, N. J., Osman, F., Maharaj, B., Naicker, N., Gibbs, A., Norman, E., Samsunder, N., Mansoor, L. E., & Karim, S. S. A. (2018). Beyond syndromic management: Opportunities for diagnosis-based treatment of sexually transmitted infections in low- and middle-income countries. PLoS Medicine, 15(4), e1002561. (10.1371/journal.pmed.1002561)

Mulu, W., Yimer, M., Zenebe, Y., & Abera, B. (2017). Common causes of vaginal infections and antimicrobial susceptibility pattern of aerobic bacterial isolates in women of reproductive age attending at Felegehiwot referral Hospital, Ethiopia: A cross sectional study. BMC Women’s Health, 17(1), 42. (10.1186/s12905-017-0397-y)

Odoki, M., Aliero, A. A., Tibyangye, J., Maniga, J. N., Wampande, E., Kato, C. D., Agwu, E., & Abigaba, G. (2019). Prevalence of bacterial urinary tract infections and associated factors among patients attending hospitals in Bushenyi district, Uganda. International Journal of Microbiology, 2019, 4246703. (10.1155/2019/4246703)

Tadesse, B. T., Ashley, E. A., Ongarello, S., Havumaki, J., Huber, M., Harris, S. R., & Dittrich, S. (2017). Antimicrobial resistance in Africa: A systematic review. BMC Infectious Diseases, 17(1), 616. (10.1186/s12879-017-2713-1)

Unemo, M., Lahra, M. M., Cole, M., Galarza, P., Ndowa, F., Martin, I., Dillon, J. R., Someshwar, M., George, R., & Wi, T. (2019). World Health Organization Global Gonococcal Antimicrobial Surveillance Program (WHO GASP): Review of new data and evidence to inform international collaborative actions and research agendas. Sexually Transmitted Infections, 95(8), 587–593. (10.1136/sextrans-2019-054043)

World Health Organization. (2016). Global health sector strategy on sexually transmitted infections 2016-2021: Toward ending STIs. World Health Organization. (https://apps.who.int/iris/handle/10665/246296)

